# Mental health in higher-education students and non-students: evidence from a nationally representative panel study

**DOI:** 10.1101/2020.09.18.20196782

**Authors:** Evangeline Tabor, Praveetha Patalay, David Bann

## Abstract

Despite increasing policy focus on mental health provision for higher education students, it is unclear whether they have worse mental health outcomes than their non-student peers. In a nationally-representative UK study spanning 2010-2019 (N=11,519), 17-24 year olds who attended higher education had lower average psychological distress (GHQ score difference=–0.37, 95%CI: −0.60, −0.08) and lower odds of case-level distress than those who did not (OR=0.91, 95%CI: 0.81, 1.02). Increases in distress between 2010 and 2019 were similar in both groups. Accessible mental health support outside higher education settings is necessary to prevent further widening of socioeconomic inequalities in mental health.

## Introduction

Three-quarters of all lifetime mental disorders emerge before the age of 25, and young people are increasingly likely to report mental ill-health (1, 2). For example, one analysis of nationally-representative UK health surveys found the prevalence of mental health conditions increased 8% per year between 1995 and 2014 (2). In particular, young women appear to experience worse outcomes than their male peers (2). Approximately half of young people currently attend a higher education institution in the UK, which combined with concern around mental health among students has prompted recent focus on university mental health provision (3, 4). However, despite these worsening trends overall, it is unclear if those attending higher education—a comparatively socioeconomically advantaged group (3)—experience worse mental health than their non-student peers (5, 6). Understanding such differences is important to inform the allocation of resources to improve population mental health and better understand the causes of population-level mental health change. However, to our knowledge there are no large nationally-representative studies in the UK which have addressed this question. Existing studies have relied on convenience samples (7) and/or used small sample sizes (e.g. N<200 (5)). In the current study we present data comparing higher-education students and non-students among 17-24 year olds from a large nationally-representative household panel study and examine trends in this difference from 2010 to 2019.

## Methods

We used data from eight waves of Understanding Society: the UK Household Longitudinal Study (UKHLS), collected between 2010 and 2019. UKHLS is a longitudinal panel survey of approximately 40,000 households across England, Scotland, Wales and Northern Ireland which started in 2009(8). Data collected in the first wave from 2009-2010 was excluded as comparable data for key variables and controls were not available. Further information concerning the sample design and measures are available elsewhere(8). The University of Essex Ethics Committee approved all data collection conducted as part of the UKHLS main study.

We selected respondents aged 17-24 years who had valid data for sex, ethnicity, parental education qualification, and mental health variables. To ensure our analysis was not biased by the exclusion of those with missing data for those variables, we conducted a sensitivity analysis on the sample restricted only by age and self-completion questionnaire response (findings were unchanged). Respondents who were attending a university or a higher or further education college or had a degree were included in the higher education sample (43.9%). Those in employment, apprenticeships/other training, not in employment or training and who did not have a degree were included in the non-higher education sample (56.1%).

Mental health outcomes were measured using the General Health Questionnaire (GHQ-12). The GHQ-12 is a validated measure of psychological distress and responses on 12 items are summed resulting in a score ranging from 0 to 36 (higher scores indicating more distress)(9). Probable psychiatric caseness was assessed as an additional outcome, with scores of 12 or more indicating an outcome consistent with a diagnosable common mental disorder (9).

Associations between student status and outcomes were examined using both linear regression (continuous GHQ scores) and logistic regression (binary caseness). Analyses were unadjusted, then adjusted for multiple possible confounders (sex, age, ethnicity, and highest parent educational qualification). We conducted analyses in a model pooled across all years using the observations at the midpoint of each unique participant’s involvement in the panel study, and separately in each year to examine time trends. We also performed an analysis separately in males and females to elucidate any differences by student status and sex. All analyses accounted for the complex survey design of the study and the appropriate non-response weights. Analyses were conducted with STATA v15.1. Further methodological information is available in the Supplementary Material.

## Results

For the pooled analyses across years we analysed data from 11,519 participants (43.9% higher education students). The sample size of those aged 17-24 was 4,404 in 2010-11 to 3,277 in 2017-19 (Table S1). The proportion of the sample categorized in the higher education group was higher in 2017-19 (48.1%) compared with 2010-11 (42.1%). Higher education status was associated with being older, female, White British, and having higher parental education (Table S2).

Across all years, those who attended higher education had lower GHQ scores than those who did not (−0.36 (−0.65, −0.08 95% CI) in unadjusted models, and −0.37 (−0.66, −0.08 95% CI) after adjustment) (Fig. 1); Cohen’s d=-0.80. This direction of association was the same in all but one year, with strongest evidence in 2010-11 and 2015-17 (Fig. 1, Table S3).

**Figure 1:**
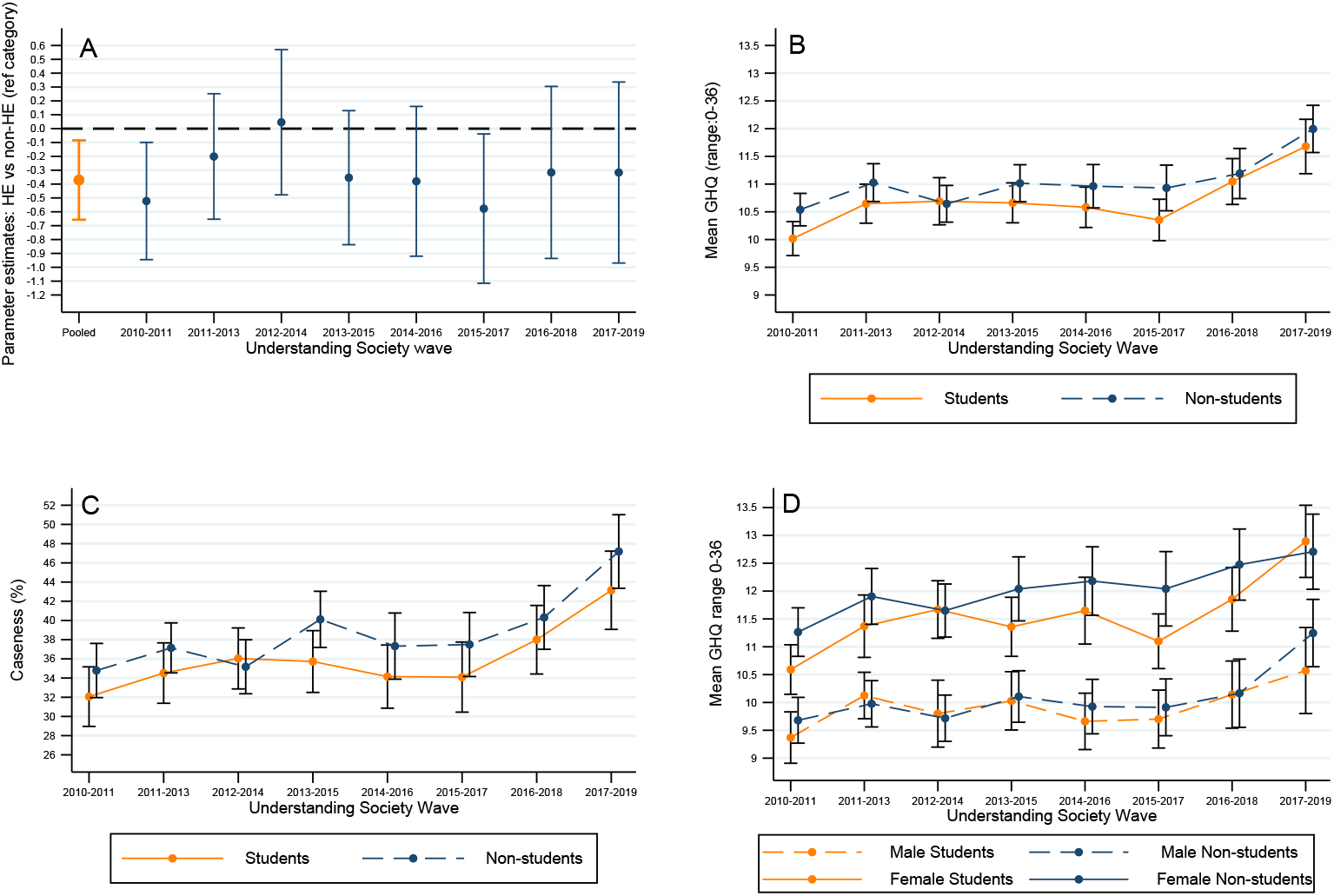
Mental health in higher education students compared with non-students: (A) regression estimates in pooled and repeated cross-sectional analyses. (B) means by study year (C) psychological distress prevalence by study year (D) means by study year and gender. Notes: all estimates adjusted for age, ethnicity, parents’ education; A-C adjusted for gender. Error bars represent 95% confidence intervals

Findings were broadly similar when using binary caseness outcome, yet as anticipated associations were less precisely estimated likely due to loss of information due to binarization (Fig, 1, Table S4): odds ratio of caseness for the higher education group against the non-higher education group: 0.91 (0.81, 1.02 95% CI) after adjustment. The lower GHQ scores amongst the higher education group was also found in both genders (Table S3-4). Amongst both education groups—and in both sexes—GHQ scores were higher in later years (Fig. 1).

## Discussion

Using nationally representative data from years 2010 - 2019 we found that overall the higher education group had better mental health than the non-higher education group. This result persisted when analyses were adjusted for sex, ethnicity, and highest parental educational qualification, and when conducted separately by gender. In addition, both groups saw substantial worsening of psychological distress outcomes between 2010 and 2019. Our findings, highlighting the value of large nationally representative surveys, are contrary to previous work in online convenience and smaller population samples which suggested those in higher education had similar or worse mental health outcomes than their non-attending peers (5, 7).

There are multiple possible explanations for our findings. First, higher education students are comparatively socioeconomically advantaged—this in turn is associated with better mental health outcomes (3, 10). Thus, our findings may capture unmeasured socioeconomic differences between the two groups. Conversely, young people with pre-existing mental health concerns are less likely to attend higher education and are at greater risk of attrition (6). In addition, higher education often confers access to resources—such as fulfilling work and new social opportunities—which have beneficial outcomes for mental health (10).

A limitation of the study was the higher education category does not distinguish between type of higher education institution, nor the type or level of qualification being sought. Additionally, the study was not powered to detect small differences between groups across time and hence annual time trends in differences between those attending and not attending higher education are difficult to establish with confidence. Lastly, the time period of focus here includes the introduction of higher tuition fees in some countries in the UK, however, sub-samples within each country, especially Scotland and Northern Ireland, were too small to investigate the differential trends by country.

Our findings indicate that despite increased focus on universities and other higher education providers to improve student mental health support, resources and attention should not be uniquely focused on the higher education population(4). Focussing largely on higher education settings to provide mental health support for this age group, despite their many advantages as sites of intervention, may lead to the inadvertent widening of socioeconomic inequalities in mental health (4, 10).

## Data Availability

The UKHLS data are available to all researchers, free of cost from the UK Data Service.

https://www.ukdataservice.ac.uk

## Acknowledgements

Understanding Society is an initiative funded by the Economic and Social Research Council and various Government Departments, with scientific leadership by the Institute for Social and Economic Research, University of Essex, and survey delivery by NatCen Social Research and Kantar Public. The research data are distributed by the UK Data Service.

## Author Contribution

ET conducted all the analysis and led on drafting the manuscript. DB and PP conceptualised the study and supervised data analysis, interpretation and drafting of the manuscript. All authors approve the final version and consented to its publication.

## Data Availability

The UKHLS data are available to all researchers, free of cost from the UK Data Service (https://www.ukdataservice.ac.uk).

## Ethics Statement

The University of Essex Ethics Committee has approved all data collection conducted as part of the UKHLS main survey.

## References

1. Kessler RC, Berglund P, Demler O, Jin R, Merikangas KR, Walters EE. Lifetime prevalence and age-of-onset distributions of DSM-IV disorders in the National Comorbidity Survey Replication. Arch Gen Psychiatry. 2005;62(6):593–602.

2. Pitchforth J, Fahy K, Ford T, Wolpert M, Viner RM, Hargreaves DS. Mental health and well-being trends among children and young people in the UK, 1995-2014: analysis of repeated cross-sectional national health surveys. Psychological Medicine pp 1-11 (2018). 2018.

3. HESA. Higher Education Student Statistics: UK, 2018/19 - Student numbers and characteristics. Cheltenham: Higher Education Statistics Agency; 2020.

4. Hughes G, Spanner L. The University Mental Health Charter. Leeds: Student Minds; 2019.

5. McManus S, Gunnell D. Trends in mental health, non-suicidal self-harm and suicide attempts in 16-24-year old students and non-students in England, 2000-2014. Soc Psychiatry Psychiatr Epidemiol. 2020;55(1):125–8.

6. Auerbach RP, Alonso J, Axinn WG, Cuijpers P, Ebert DD, Green JG, et al. Mental disorders among college students in the World Health Organization World Mental Health Surveys. Psychol Med. 2016;46(14):2955–70.

7. Neves J, Hillman N. Student Academic Experience Survey 2017. York: Higher Education Academy & Higher Education Policy Institute; 2017.

8. Lynn P. Sample Design for Understanding Society. Colchester: Institute for Social and Economic Research; 2009.

9. Lundin A, Hallgren M, Theobald H, Hellgren C, Torgén M. Validity of the 12-item version of the General Health Questionnaire in detecting depression in the general population. Public Health. 2016;136:66–74.

10. Montez JK, Friedman EM. Educational attainment and adult health: Under what conditions is the association causal? Social Science & Medicine. 2015;127:1–7.

